# COVID-19 vaccination at a hospital in Paris: spatial analyses and inverse equity hypothesis

**DOI:** 10.1101/2023.05.05.23289561

**Authors:** Ridde Valéry, André Gaëlle, Bouchaud Olivier, Bonnet Emmanuel

**Affiliations:** Université Paris Cité, IRD, Inserm, Ceped, F-75006 Paris, France; Master Carthagéo, University Paris 1 Panthéon-Sorbonne, UMR 215 Prodig, IRD, CNRS, AgroParisTech, 5, course of Humanities, F-93 322 Aubervilliers Cedex, France; Hospital Avicenne-Assistance Publique hospitals de Paris and Université Sorbonne Paris Nord, F-93000 Bobigny, France; IRD, UMR 215 Prodig, CNRS, Université Paris 1 Panthéon-Sorbonne, AgroParisTech, 5, course of Humanities, F-93 322 Aubervilliers Cedex, France

**Keywords:** vaccination, COVID-19, spatial analysis, equity, mediation, France

## Abstract

**Background:** Vaccination against SARS-CoV-2 has been deployed in France since January 2021. Evidence was beginning to show that the most vulnerable populations were the most affected by COVID-19. Without specific action for different population subgroups, the inverse equity hypothesis postulates that people in the least deprived neighbourhoods will be the first to benefit.

**Methods:** We performed a spatial analysis using primary data from the vaccination centre of the Avicenne Hospital in Bobigny (Seine-Saint-Denis, France) from January 8th to September 30th, 2021. We used secondary data to calculate the social deprivation index. We performed flow analysis, k-means aggregation, and mapping.

**Results:** During the period, 32,712 people were vaccinated at the study centre. Vaccination flow to the hospital shows that people living in the least disadvantaged areas were the first to be vaccinated. The number of people immunized according to the level of social deprivation then scales out with slightly more access to the vaccination centre for the most advantaged. The furthest have travelled more than 100 kilometres, and more than 1h45 of transport time to get to this vaccination centre. Access times are, on average, 50 minutes in February to 30 minutes in May 2021.

**Conclusion:** The study confirms the inverse equity hypothesis and shows that vaccination preparedness strategies must take equity issues into account. Public health interventions should be implemented according to proportionate universalism and use community health, health mediation, and outreach activities for more equity.

## Introduction

In February 2021, at the heart of the COVID-19 pandemic, The Lancet recalled the importance and relevance of Julian Tudor Hart’s 1971 proposal: the inverse care law (1). While Tudor’s proposal was initially applied to healthcare, it also adapts to public health interventions and the unfair distribution of health outcomes. This phenomenon is so common that Watt asserted *“the monstrous longevity of the inverse care law”* (2). Social justice and equity issues have always been at the heart of public health and vaccinations. Therefore, action to reduce social inequalities in health remains a daily challenge for public health organizations, including in France (3). Aiming to achieve effectiveness at the expense of equity is an ongoing challenge for people planning and implementing public health interventions such as vaccinations (4). Most health phenomena confirm this inverse care law, including the distribution of SARS-COV-2 in most countries, even though data collection is still ongoing (5,6). In France, research on the first wave showed significant spatial heterogeneity in hospital COVID-19 incidence and mortality distribution (7,8). The evidence demonstrated that from the onset of the pandemic, the most vulnerable and precarious populations were most affected—especially people born abroad and living in vulnerable neighbourhoods (9–11).

The corollary to Hart’s proposals is the importance of interventions to address this uneven distribution: the famous principle of Marmot’s proportional universalism (12). Numerous articles, books, guides for action, and public health conferences in France have raised this challenge of equity and the need for it to be considered by public health organizations. Public health officials in the Ile de France region, where Paris and this present study are located, were fully aware of these challenges at the beginning of the pandemic. They had engaged in reflections to consider them even though vaccination was not yet on the agenda at that time (13).

In France, as elsewhere in the world, considering the inverse care law when planning public health interventions is neither obvious nor a reflex. Reviews show that people planning infectious disease interventions need to be more explicitly concerned about the issues of equity (14,15). These challenges were identified, for example, in Paris (France) for the same follow-up and the organization of SARS-CoV-2 tests organized by the hospital network (16). However, other arrangements have been organized by the Regional Health Agency (ARS) to promote regional equity, beyond these hospital facilities, without conclusive results (17).

At the end of 2020, vaccination against SARS-CoV-2 became the world’s leading public health intervention, and experts explained the challenges of inequality it imposed (18). The first studies on vaccination in Israel confirm the socio-economic gradient (19). Yet empirical studies on inequalities and vaccination against SARS-CoV-2 remain rare (20). Vaccine inequalities are well-known in France for other antigens (21).

Initially focused on health professionals and vulnerable populations in France, a free-of-charge vaccination campaign was gradually proposed to all. The main dates for the deployment of the vaccination strategy are listed in Appendix 1. From January 15th, 2021, appointments in vaccination centres were made by telephone and Internet without a medical prescription, but people could also come to the centres without an appointment. The national strategy was initially designed regarding biomedical risk factors without considering social vulnerabilities. The structures closer to the population (without appointments: pharmacies, liberal medicine, etc.) were only deployed later, especially according to the demands of the field actors.

Seine-Saint-Denis, a territory particularly affected by the pandemic in the Ile de France region, and marked by significant social inequalities in historical health (13), set up vaccination centres for the general population in several hospitals in the public network and other non-hospital sites in the region (e.g. Stade de France). Given the specific needs of that department and contrary to national doctrine, the ARS had already decided to allocate more doses than a *“simple population distribution”* (22). The objective of these centres nested in the hospital network was to facilitate access to vaccination for local populations by endorsing them to hospital facilities identified and known to the population. At the COVID-19 centre of the hospital concerned by this study, after proposing a contact case screening and follow-up activity (16), vaccinations were organized from January 2021 onward. Previous qualitative research has shown that this centre’s managers and stakeholders could not consider the challenges of social health inequalities in the screening and follow-up of contact cases of SARS-CoV-2 (16). Thus, this article aims to study with quantitative data whether the inverse care law has been verified in the context of vaccination against SARS-CoV-2 from a vaccination centre of a hospital in the Paris region.

## Methods and Materials

### Hypothesis

Given the scientific knowledge, the local and hospital context of this vaccine strategy (16), and the fact that vaccination against SARS-CoV-2 is now open to the entire population without any particular restrictions, we hypothesize that people living in the least disadvantaged neighbourhoods were the first to benefit from it.

### Study period

The study is based on primary vaccination data from the vaccination centre of the hospital Avicenne AP-HP (Assistance Public Hopitaux de Paris) in Bobigny in Seine-Saint-Denis from January 8th to September 30^th^, 2021.

### Data source

The vaccination database was obtained from the ORBIS® application. It is pseudonymized and contains variables on the vaccinated person’s vaccination date, age, and postal address. To produce maps, we used secondary data to calculate access times between residential neighbourhoods (INSEE’s IRIS — Institut National de la Statistics et des Etudes Economiques) and the vaccination centre. Those data come from the “open data Ile de France Mobility”. To calculate the Social Deprivation Index, we worked from the FDEP index (Social Disadvantage Index of the National Institute of Health and Medical Research (Inserm).

### Data aggregation

All vaccination information at the address has been aggregated to the IRIS reference. IRIS is a basic geographical unit for disseminating infra-communal data in France. It contains between 1800 and 5000 inhabitants. We then disaggregated these data into a grid composed of 800m^2^ tiles and mapped the values of the vaccination data. This grid mapping method makes it possible to use different levels of aggregation and disaggregation of variables by merging spatial information plans, here address, IRIS and tile. It also preserves the anonymity of vaccinated people.

### Spatial and statistical analysis

To assess vaccinated people’s residential origin, we analyzed the vaccination flows (flow matrix) between the residence IRIS and the vaccination centre. We then mapped these flows to connect the residence and the place of vaccination. To integrate flow intensity, k-means aggregation creates spatialized clusters of vaccination flows. Secondary data were used to calculate access times between IRIS and the vaccination centre based on Ile de France’s road transport networks and public transport networks.

As regards the inclusion of vulnerable persons, we used the French Deprivation Index (FDep), which produces a geographical indicator of the general population of the social disadvantage. The FDep combines material and social disadvantages on the geographic scale of IRIS. It makes it possible to highlight a dimension that maximizes spatial variability at the socio-economic level. Its calculation is carried out with a population-weighted primary component analysis based on four variables from INSEE (unemployment rate in the labour force, labour rate in the labour force, bachelor rate in the out-of-school population over 15 years of age, and the median income reported per unit of consumption) made it possible to carry out this analysis. The association of the deprivation index (Appendix 2) and the matrix allow a bivariate flow mapping representing the original flows and their level of social deprivation.

The Committee for Evaluating the Ethics of Biomedical Research Projects (CEERB) Paris Nord (IRB: 00006477) has reviewed and approved the research. No nominative data were used. The analyses were presented and validated by staff involved in the vaccination campaign.

## Results

During the period, 32712 people were vaccinated in the study centre. The number of people vaccinated (Figure 1) first evolved with the different phases of the COVID-19 vaccination program. According to the national policy, health workers benefited from it first, then biologically vulnerable people over 50, then people over 70, and then people over 50 between March and May 2021. From May 18, 2021, vaccinations were open to all people 18 and older. At the beginning of the campaign, people living in the most favoured neighbourhoods had the most access to vaccinations. The number of people vaccinated according to the level of social deprivation then balances with a little more access to the vaccination centre for the most favoured.

**Fig 1:**
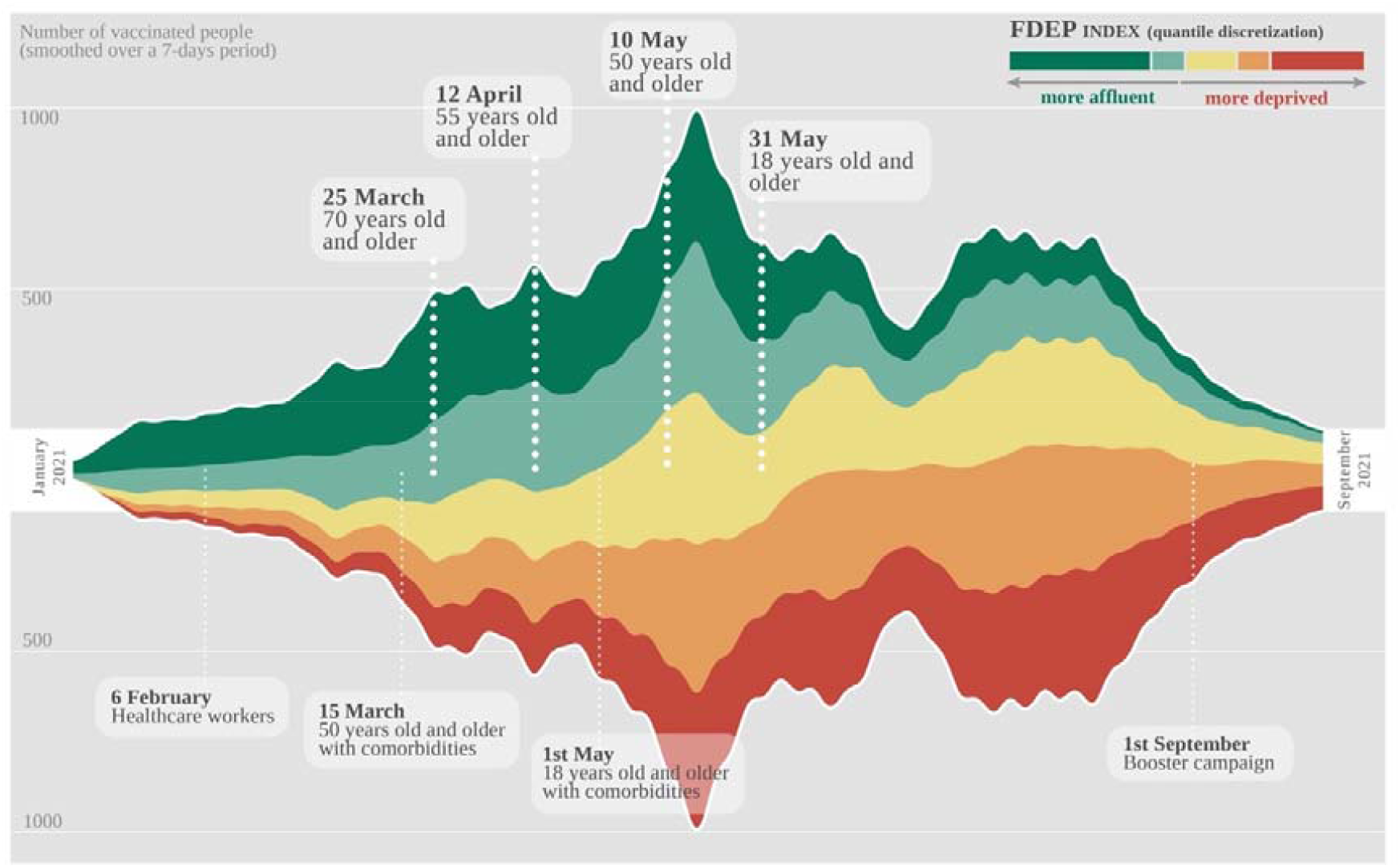
A timeline of the COVID-19 vaccination phase according to the FDED index

The other results confirm the assumption of the inverse care law (Figure 2). Vaccination flow to the hospital shows that people living in the least disadvantaged areas were the first to be vaccinated in the first months of 2021. Figure 2 shows the number of people vaccinated in the study centre by residential tiles. In January 2021, the geographical origin of the first vaccination at the hospital came from all Parisian districts and other departments of Ile de France. If it is first (in January), health workers who potentially work at the Avicenne Hospital (study centre) and reside in Paris. Without considering these personnel, this trend continues in the following months only for those affected by the vaccination phases. It was in April 2021 that the number of vaccinated people living near the hospital intensified. The area of attraction remains very important, although other centres have opened in the different parts of the territory of Ile de France. The furthest have travelled more than 100 kilometres and more than 1h45 of transport time to get to this vaccination centre (Table 1). The most people vaccinated were those closest to the vaccination centre. Access times are, on average, 30 minutes from May compared to 50 minutes in February (Appendix 3).

**Table 1:**
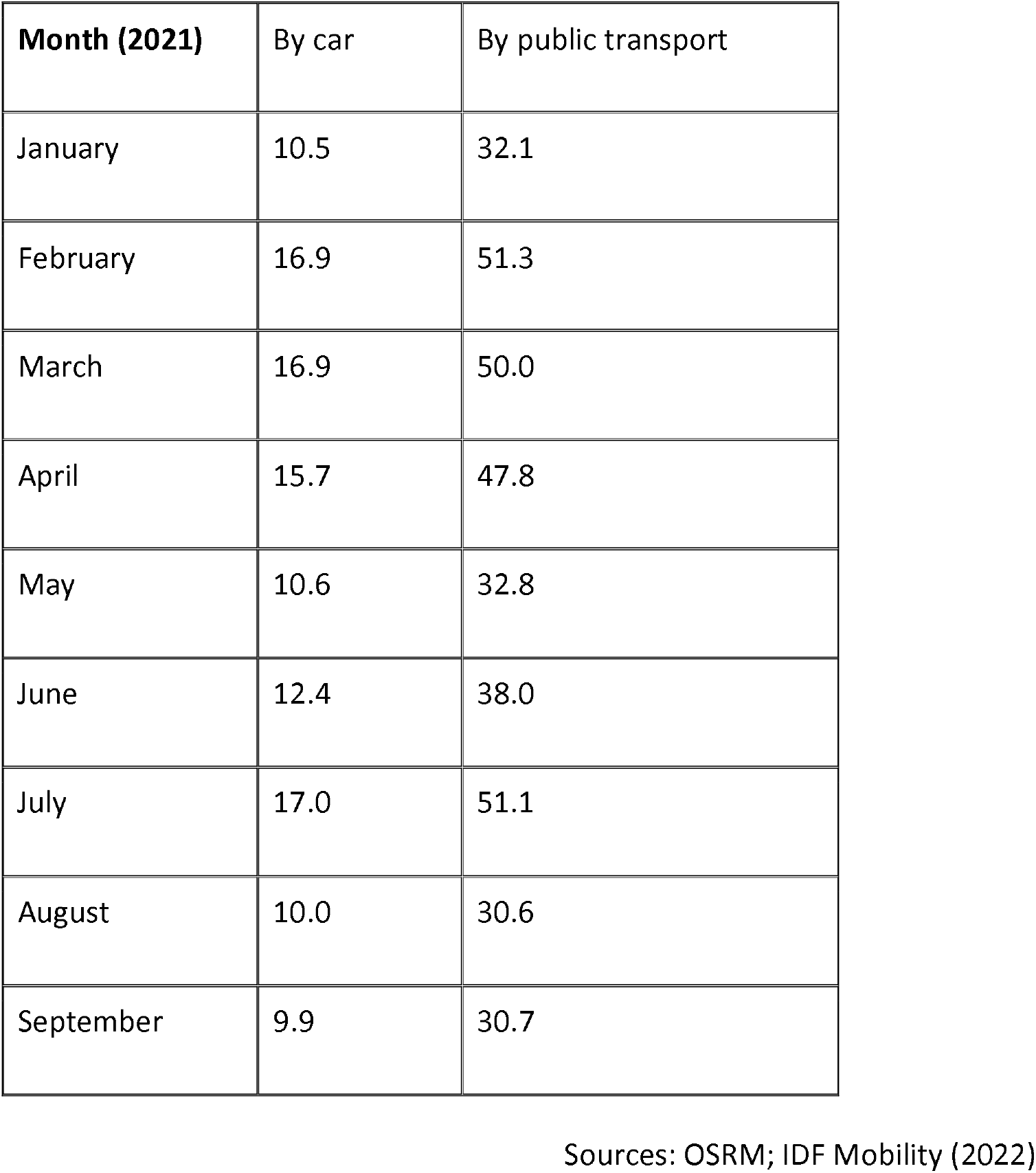
Average time distance (in minutes) travelled by people vaccinated at study centre

| **Month (2021)** | By car | By public transport |
| --- | --- | --- |
| January | 10.5 | 32.1 |
| February | 16.9 | 51.3 |
| March | 16.9 | 50.0 |
| April | 15.7 | 47.8 |
| May | 10.6 | 32.8 |
| June | 12.4 | 38.0 |
| July | 17.0 | 51.1 |
| August | 10.0 | 30.6 |
| September | 9.9 | 30.7 |

Sources: OSRM; IDF Mobility (2022)

**Figure 2:**
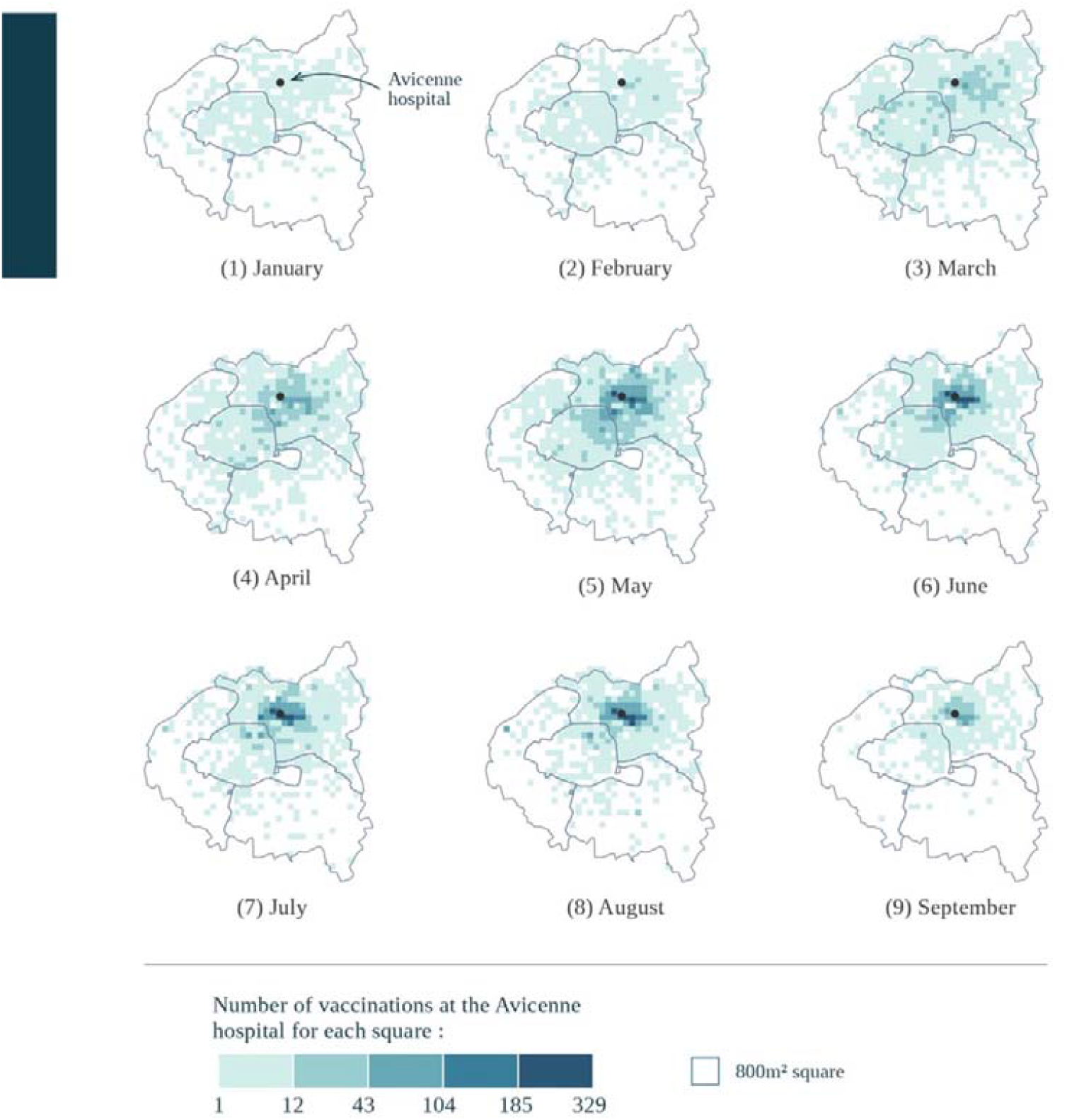
Number of COVID-19 vaccinations per residence tile

According to the Deprivation Index (Figure 3), the evolution of the vaccination flow to the hospital shows that people living in the least disadvantaged areas were the first to travel to be vaccinated in the first months of 2021. From May 2021 onwards, people from the most underprivileged areas became the majority. In addition, the proportion of people coming from less-favoured regions increases over the months and the opening of vaccination to a broader audience.

**Figure 3:**
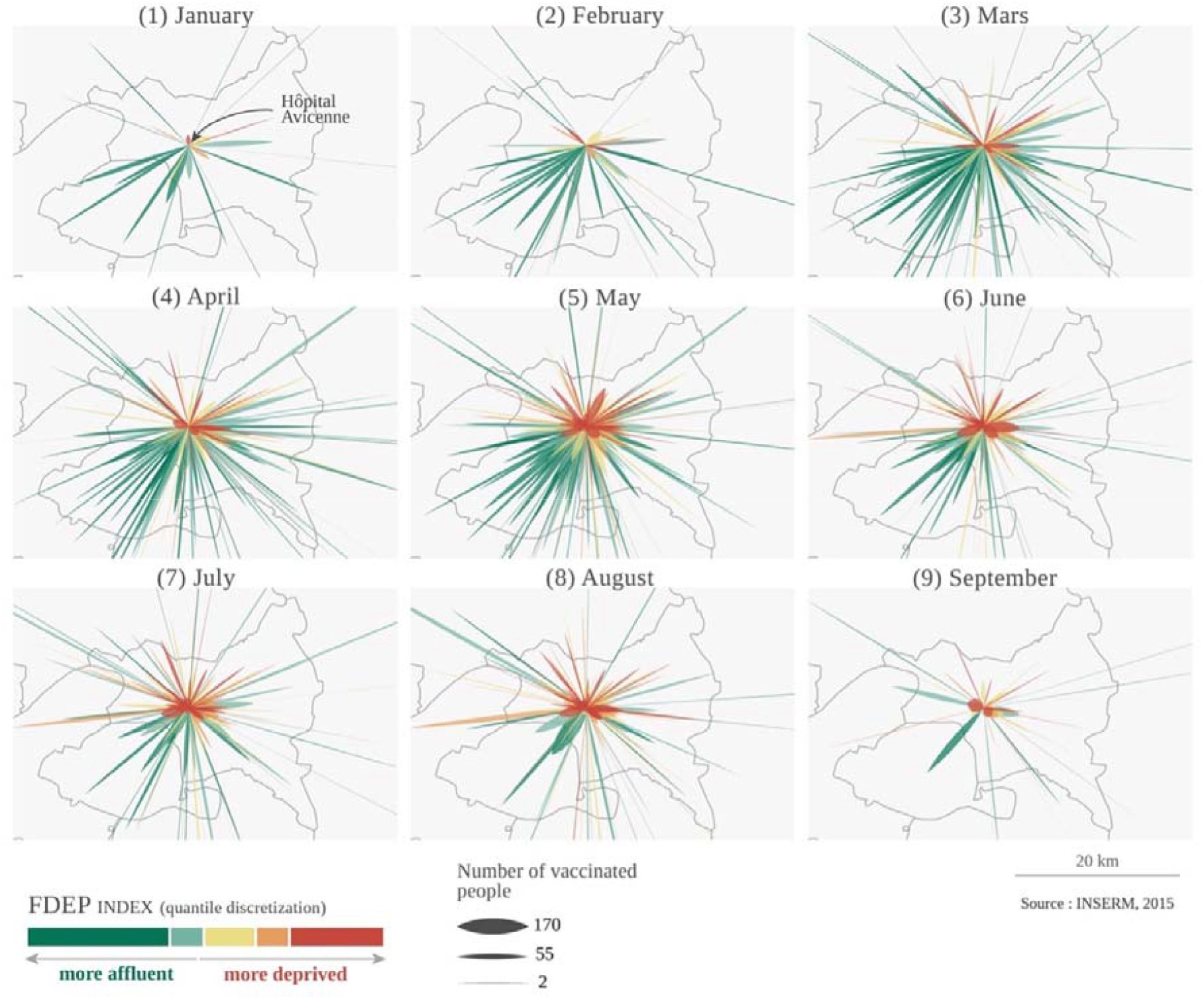
Origin of vaccinated people at the study centre in 2021

## Discussion

The results confirm that the inverse equity hypothesis once again applied to the COVID-19 vaccination campaign from a hospital in the Paris region located in a socially disadvantaged region. The centre’s opening, without geographical restriction, certainly explained the vast attraction area and inverse care law. People living in the favoured neighbourhoods of Paris used the vaccination centre more than the population of the underprivileged districts of Seine Saint Denis. The over-doing of vaccines was not effective initially because it did not reach the local population. This new pandemic confirms the need to address equity issues better when organizing vaccination campaigns by not limiting itself first to questions of care offerings but by acting on all the social determinants of vaccination, especially on the demand side.

Without going back to the old historical analyses of the lack of consideration for social inequalities in health in France and in particular in public health (23,24), contemporary history confirms its permanence on the scale of an intervention organized by the regional branch of the Ministry of Health (ARS) in a public hospital. The COVID-19 pandemic was, therefore, no exception, and all recent studies of its manifestation in France show that the most socially vulnerable people were most affected and least joined, at least at first, by public health interventions (9,25–27). A study for the entire Ile-de-France region where the hospital is located confirms this equity challenge for vaccination (28). Studies concerning SARS-CoV-2 tests (29) (and thus surveillance of the epidemic and subsequent deployment of actions) and vaccination (30) in France’s second-largest city (Marseille) show that this phenomenon is not only located in the capital region. Therefore, the scale of the challenges of considering inequalities seems national.

This result could partly be explained by the national functioning of public health in France (research as intervention), its centralization, the culture of its institutions and staff need to be more trained and equipped about equity and the lack of preparedness to face such challenges (31). The mid-level management (Ministry of Health and the Directorate-General for Health) conceived the national vaccine strategy, and the national bodies of public health experts were not involved. The political stakes were central to the strategy’s formulation stage, as with the other actions to combat the pandemic (32). The evaluation of the National Public Health Strategy (2018-2022) notes that the issue of inequalities is challenging to translate into *“concrete instruments”* and notes *“the prioritisation of public policy interventions in favour of the ‘general population”* (33), which is therefore universal and far from proportional universalism (12). An evaluation about Santé Publique France states that this state agency (independent of the government) *“does not identify local actions and the approach remains essentially top-down”* (31). In addition, public health remains a very marginal activity of the health sector in France. University public health, which is often mostly a hospital level, is not open to people without a medical degree, limiting interdisciplinarity and considering the social determinants of health (34). The pandemic has allowed clinicians and health workers working in hospitals to understand public health issues better [37]. However, this may not have been enough to raise awareness of equity issues in a population-based approach. Moreover, as the vaccine policy was highly centralized, even though the hospital staff at the study centre were aware of these issues, they did not have the means or opportunities to readjust and adapt to local contexts without involving them in decision-making.

However, the public administration anticipated these challenges at the Île de France region (Paris) level and prepared itself accordingly (17). Before vaccination, many outreach strategies were organized for screening (35). Then, an additional allocation of vaccines was granted for the department where the hospital is located (in a national context where the doctrine was to lose no dose), and dozens of “mediators” (anti-COVID) worked in the territories between June and December 2021. The national health insurance staff called on the phone to older people or in precarious situations to suggest they make an appointment for a vaccination. In May 2021, a national workshop, with the presence of people from the Ile-de-France region, was organized by Santé Publique France to reflect on the challenges of vaccination of people in precarious situations (35). In particular, they advocated developing interventions capable of considering the needs of people in situations of great precariousness and implementing interventions in outward mode, doing with and doing together. Despite these regional public health actions and the deployment of financial resources like never before, our centre-wide analysis at a university hospital shows that this has not been enough to avoid the inverse care law. We do not have data on a scale as fine as ours to verify that this may have been different elsewhere in the region and vaccination centres deployed outside a hospital facility. However, a study in the Paris region and Marseilles shows that people in precarious situations benefited less from primary vaccination than the general population and had a two-month lag to access it (27).

At the more local level, significant funding has been offered to local health intervention organizations and associations to implement mobile and outreach strategies (22). But they have not been able to meet all the demands, undoubtedly because of the lack of commitment of the State to the sustainability of these mediation strategies, the recurrent and historical challenges of staff and the capacity to absorb the resources of local associations of a level never seen in contemporary history and finally a lack of national recognition of these health promotion approaches. However, the examples of Marseille in France or London in England show that this decentralized, participatory and inclusive approach can help enhance the fairness of vaccination campaigns against SARS-CoV 2 (30). In Marseille, *“COVID-19 mediators in the northern districts also slowly multiplied vaccination awareness and facilitated activities in pop-up sites in collaboration with local health professionals and public structures”* (30). Health mediation and community health are still underdeveloped in France, while their effectiveness in improving the equity of public health interventions has long been known (36,37).

This study shows that people in the least affluent neighbourhoods eventually benefited from vaccinations after some time. Even if it was possible to come without an appointment, it needs to be clarified that working-class neighbourhoods are aware of this provision countering the challenge of digitalization, not to mention all the other determinants of the non-use of care. The deployment of SARS-Cov-2 screenings in the poorest municipalities, long before vaccination, needed to be increased to influence the immunization of people living in peripheral neighbourhoods; as was noted in Marseilles, digital equity required to be sufficiently taken into account since appointments had to be made mainly on an internet platform (30). It would be a question of carrying out a more qualitative study in our region to describe and understand the role of the various actions undertaken as they are deployed in the neighbourhoods concerned. In the department, several charitable organizations carried out activities on demand and with funding from the public administration (22). But a priori, they needed to be of more magnitude and intensity, gradually but may be late (from June) in the campaign (22,28) and certainly face coordination challenges. Indeed, the actors at the heart of regional processes confirm that *“the results are fragmentary, sometimes uncertain, or too late”* (17). An analysis by the ARS affirms that *“the various outward actions are of insufficient scale for their effects to be visible”* (28). A qualitative study in one of the municipalities concerned shows that *“actions upstream of the operation, of the type of communication or exchange with the inhabitants, could not be optimal”* (28). The conditions for actions outreach may not have been met despite the goodwill: be together, do and do with, go to and bring to (35).

The spatial analyses used in this study are enlightening for monitoring at the local level and other epidemiological analyses (38). They highlight mobility and make it possible to cross-check data on vaccination, socio-demographic information, and distance. They produce maps that illustrate inequalities in the use of services. The geography of health associated with this type of analysis makes it possible to understand societies’ relationships with their spaces and highlight inequalities. They demonstrate the role of space in this process (39). Public health researchers and health geographers needed to be able to collaborate more often, and the disciplines better intersect. The COVID-19 pandemic illustrates that these approaches are indispensable and must be further integrated into training.

Our study has some limitations, particularly regarding the data used for January 2021. Some vaccination data could include health workers vaccinating at the study center hospital while residing in Paris. We, therefore, carried out an additional analysis by retaining only those vaccinated over 75 years of age for the first period (January 1st -January 18th 2021) to exclude health workers working in the hospital or the area (Appendix 4). This analysis confirms that the vaccinated people came from favoured neighbourhoods.

The implications of this research for public health are numerous, even if they are not necessarily very original. First, it would be helpful to translate policy statements into public health instruments better to combat social inequalities in health (33). Indeed, *“the consideration of social inequalities in the operational management of crisis remains a major task”* (17) say the public health officials of the ARS. This requires more financial resources targeted at health promotion and community health approaches. Still, more staff are trained on the operationalization of principles as soon as interventions are formulated (14,15). Tools exist, including in French, to support these approaches (40) and studies have uncovered the factors on which action could be taken to reduce vaccine inequalities related to COVID-19 (20). Clinicians, health professionals and public health personnel involved in immunization should also benefit from this equity training. Secondly, it would be helpful to go beyond hospital-centralism and the biomedical approach to health to deploy actions to promote health, community health, mediations and strategies to reach the target audiences (20,37). The sustainability of the positions of those involved in these local and participatory processes in the context of the pandemic is becoming a significant issue, which should not be forgotten in the face of the ever-greater and more media demands for funding from the curative and hospital sector. In addition, proportionate universalism should become the reference standard for our public health actions (12). Finally, public health actors should be given the means to evaluate their interventions, including vaccination and prevention, about distributing their benefits. It is, of course, very possible that many interventions have been deployed at local scales and succeeded in reducing inequalities without the availability of data to prove it without an evaluation process planned upstream (17). We can only hope that the equity issue will be at the heart of public health responses for the next epidemic.

## Data Availability

All data produced in the present work are contained in the manuscript

## Key Points

- Vaccination against SARS-CoV-2 at a hospital in Paris confirms the inverse equity hypothesis.
- People in the most affluent neighbourhoods were the first to benefit from vaccination.
- Spatial analysis methods help to understand the equity issues of public health interventions.
- Vaccination preparedness strategies must take equity issues into account.
- Considering equity when planning, implementing, and evaluating vaccination interventions is essential.

## Acknowledgements

We want to thank for their availability the people involved in the vaccination campaign, their coordinator (Jean Michel Cayrou), the hospital staff, Zoé Richard for his qualitative research; Luc Ginot, Jeanne Pergeline and Elodie Richard for their support in interpreting the results.

## Supporting information

**Appendix 1:**
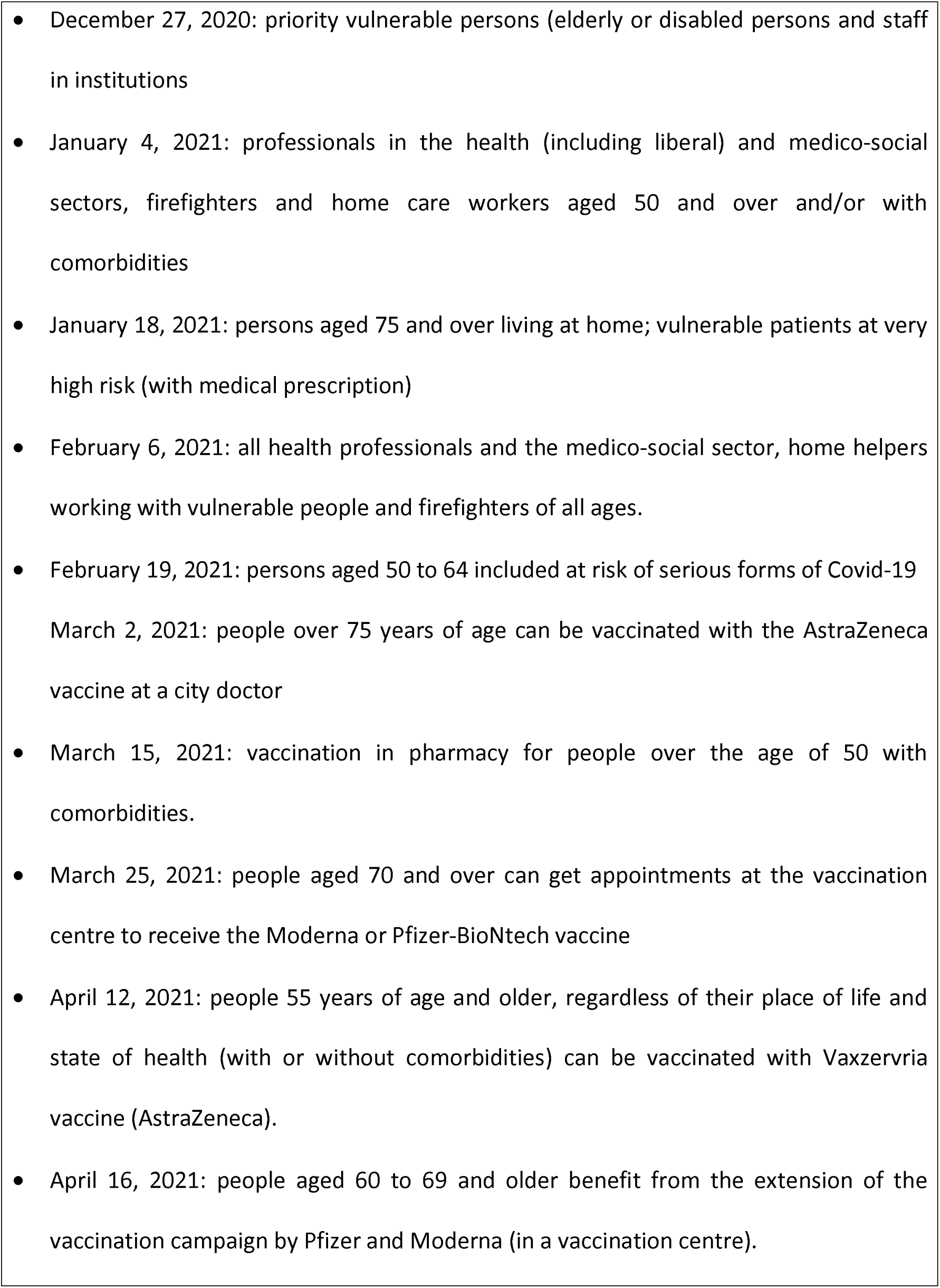
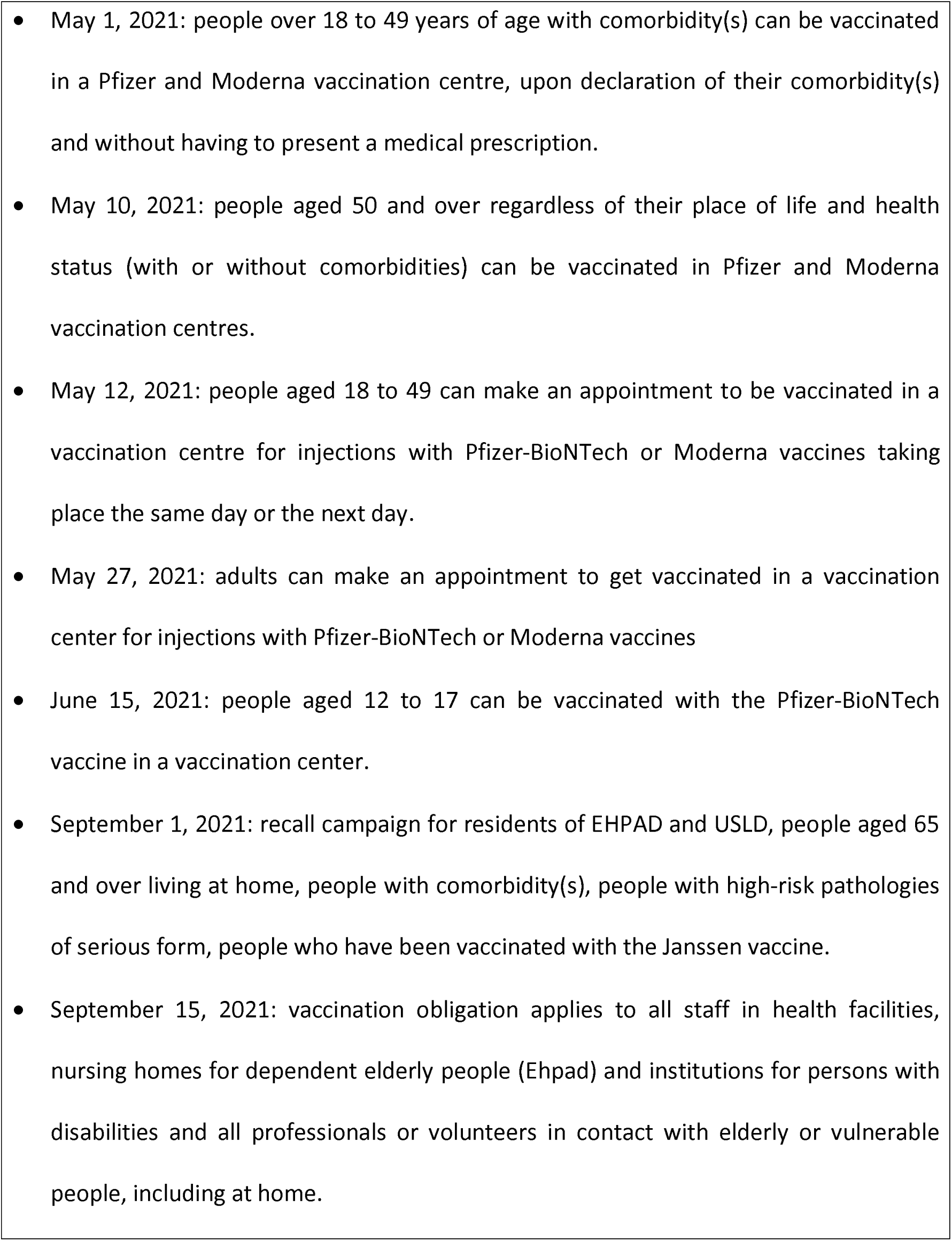
the main dates of the SARS-CoV-2 vaccine strategy in France

**Appendix 2:** Weighted averages of origin FDEP indices of vaccine at the study centre

| **Months** | Weighted average FDEP<br>for Paris (75) | Weighted average FDEP for<br>Seine-Saint-Denis (93) |
| --- | --- | --- |
| January | —1.925305 | 1.6245317 |
| February | —2.810229 | 0.7842973 |
| March | **—2.904637** | **0.7580089** |
| April | —2.337574 | 1.0021073 |
| May | —1.620894 | 1.7295278 |
| June | —2.030242 | 1.3616893 |
| July | **—2.914706** | 0.8438804 |
| August | **—1.322902** | **1.8257919** |
| September | —1.694564 | 1.7576404 |

**Appendix 3 :**
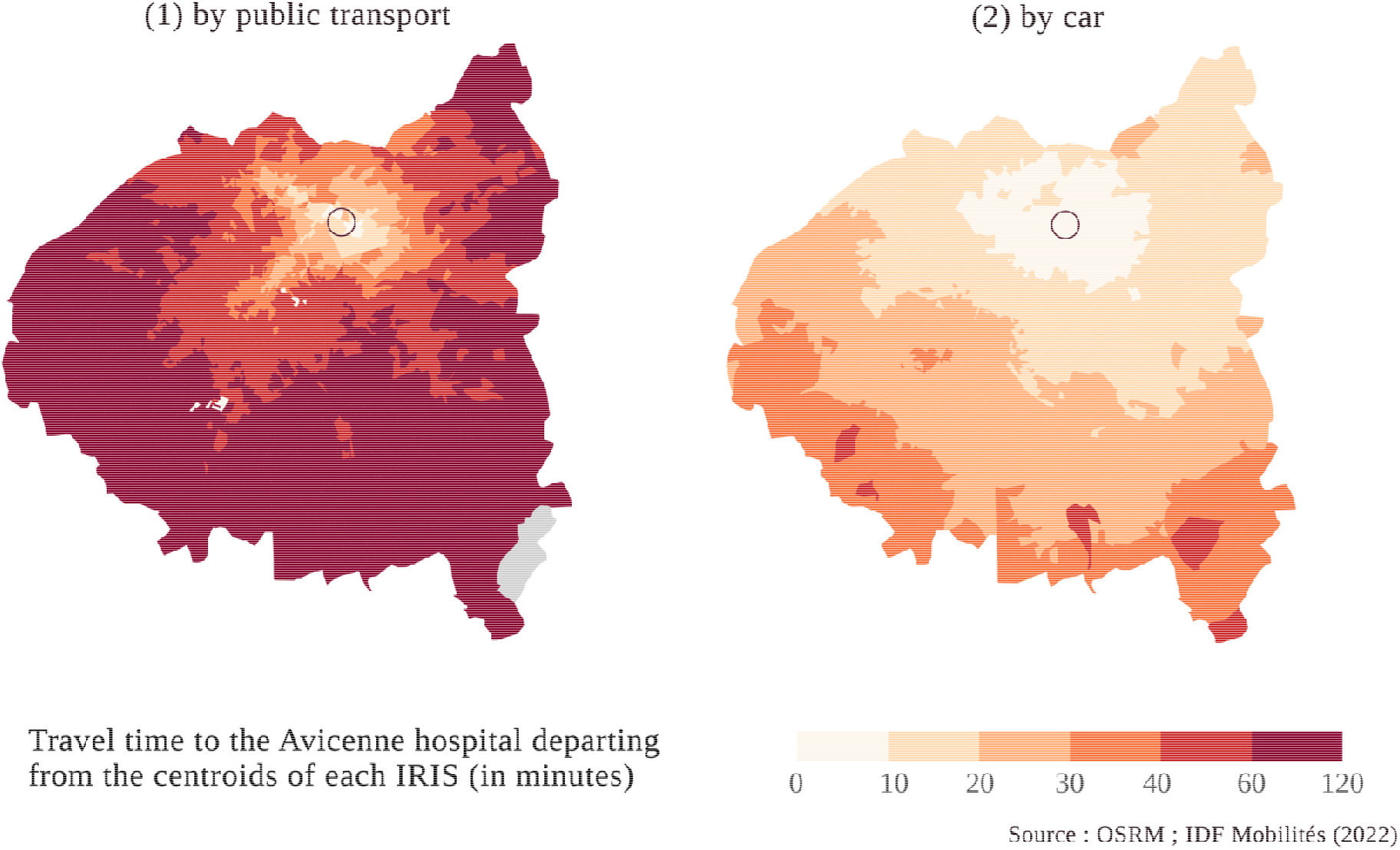
Time accessibility at the study centre

**Appendix 4:**
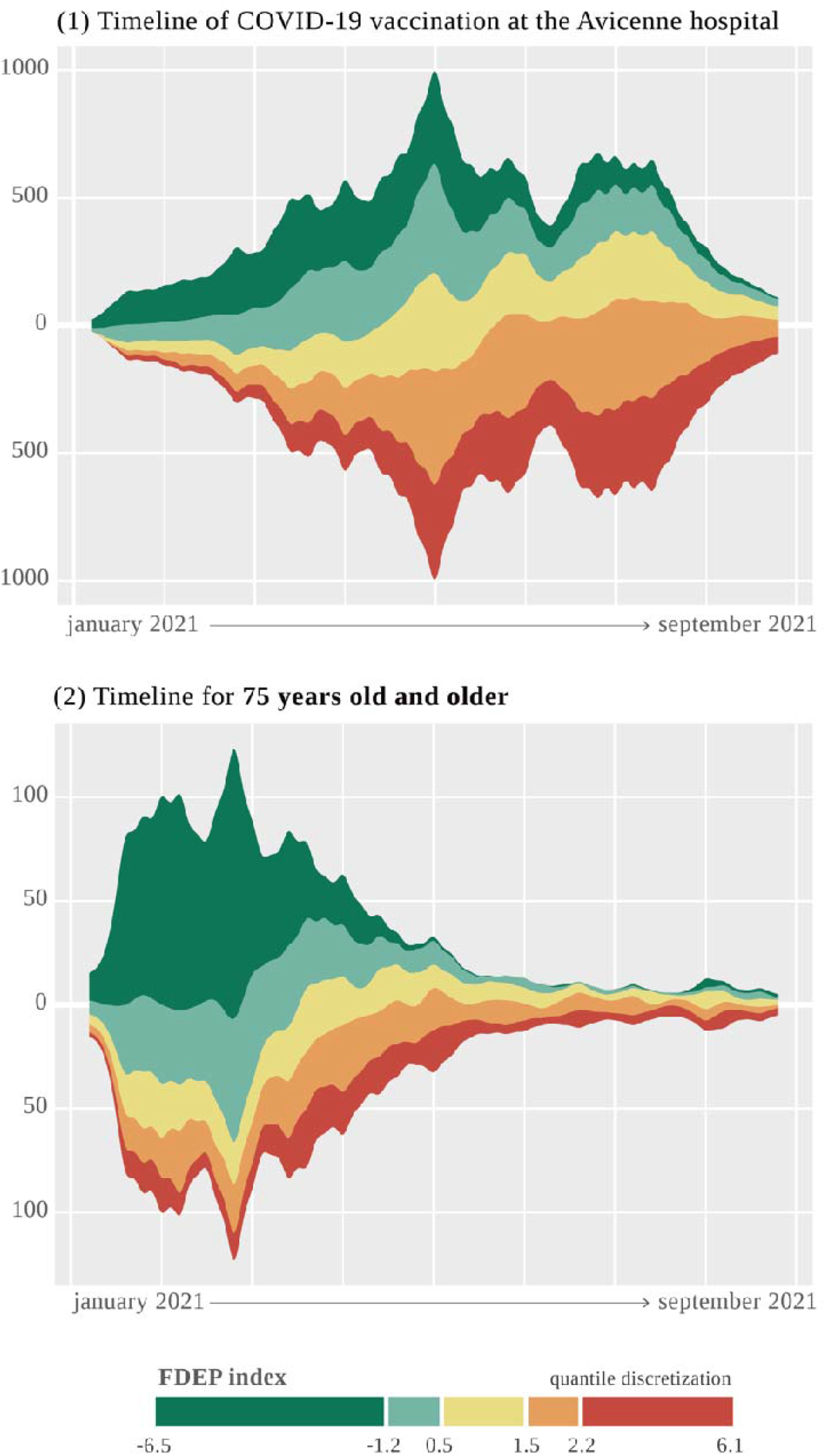
A timeline of the COVID-19 vaccination phase according to the FDED index

